# Understanding patterns of adherence to COVID-19 mitigation measures: A qualitative interview study

**DOI:** 10.1101/2020.12.11.20247528

**Authors:** Sarah Denford, Kate S Morton, Helen Lambert, Juan Zhang, Louise E Smith, G James Rubin, Shenghan Cai, Tingting Zhang, Charlotte Robin, Gemma Lasseter, Mathew Hickman, Isabel Oliver, Lucy Yardley

## Abstract

In an effort to reduce the spread of COVID-19, the UK government has introduced a series of mitigation measures. The success of these measures in preventing transmission is dependent on adherence, which is currently considered to be low. Evidence highlights the disproportionate impact of mitigation measures on individuals from Black, Asian and minority ethnic (BAME) communities, as well as among those on a low income, and an understanding of barriers to adherence in these populations is needed. In this qualitative study we examined patterns of adherence to mitigation measures and reasons underpinning these behaviors among people on low income and those from BAME communities.

Semi-structured interviews were conducted with 20 participants from BAME and low-income White backgrounds. The topic guide was designed to explore how individuals are adhering to social distancing and self-isolation measures during the COVID-19 pandemic, and to explore in detail the reasons underpinning this behavior. Data were analyzed using ‘thematic analysis following which charts were used to help compare concepts within and between participants and develop an understanding of patterns of adherence.

Participants were confused by the constantly changing and seemingly contradictory rules and guidance. As a result, decisions were made about how best to protect themselves and their household from COVID-19, and from the detrimental impact of lockdown restrictions. This was not always in line with government advice. We identified three categories of adherence to lockdown measures 1) caution motivated super-adherence 2) risk-adapted partial-adherence and 3) necessity-driven partial-adherence. Decisions about adherence considered potential for exposure to the virus, ability to reduce risk through use of protective measures, and perceived importance of/need for the behavior.

This research highlights a need for a more nuanced understanding of adherence to lockdown measures. Provision of practical and financial support could reduce the number of people who have to engage in necessity-driven partial-adherence. Information about viral transmission could help people assess the risk associated with partial-adherence more accurately. More evidence is required on population level risks of people adopting risk-adapted partial-adherence.

## Introduction

Numerous mitigation measures have been introduced in an effort to prevent the spread of coronavirus (COVID-19) in the United Kingdom (UK). These include restrictions of movement, self-isolation, social distancing, mandatory use of face-coverings in certain settings, and engagement in test, trace and isolate procedures when necessary. Critically, the effectiveness of these measures for preventing transmission is dependent on the extent to which they are known, understood, accepted and adopted in time. However, throughout the pandemic, research indicates variation in the extent to which people are willing and able to adhere to guidance (1-4).

A growing number of surveys have been conducted with the aim of identifying factors associated with adherence to lockdown, self-isolation and social distancing guidance. These surveys highlight demographic influences such as age (5), gender (3, 5, 6), and ethnicity (2). Perceptions of risk (5, 7), behavior of others (8), access to help and support (3), trust in the government and the effectiveness of mitigation measures (6, 8) and already having had COVID-19 (9) may also influence behavior. More recently, qualitative research has been used to explore in-depth experiences of adherence to lockdown measures, and reasons for non-adherence (10). Focus groups with UK residents suggested that exposure to too much, or constantly changing information, a lack of trust and respect for those in power, and resistance and rebelliousness may contribute to a lack of adherence to guidelines (10). However, much of this research has focused on predicting or explaining adherence, rather than focusing on what people are doing, why, and how safe it is.

While social distancing and self-isolation behavior during lockdown can be crudely defined as being “adherent” or “not-adherent,” it is likely that there is a more nuanced scale of adherence (10). Attempts have been made to challenge the view that adherence should be considered a dichotomy. For example, Fancourt et al use the terms “complete” and “majority” adherence to compare those who follow all the guidance all the time with those who follow some of the guidance, or for some of the time (11). Williams et al refer to “overt rule breaking” and “subjective rule interpretation” to differentiate between those who were deliberately breaking the rules, and those who are interpreting inconsistent or constantly changing guidance to suit their needs (10). However, these terms do not adequately capture the complexities underpinning decisions to adhere (or not) to the guidance, and the risk that this may bring. People may be acting conscientiously, engaging only in activities in which they are unlikely to come into contact with anyone, attempting to minimize the risk of transmitting or catching the virus (10).

Alternatively, people may be leaving their home out of necessity, in order to buy food or medicine, for a medical need, or to provide care for a vulnerable person. It is still unclear how the public are making decisions about what they should and should not do to control infection transmission, what they actually are doing, and how safe this is.

Periods of lockdown may be particularly challenging for those from lowest income backgrounds and individuals from black Asian and minority ethnic (BAME) communities who are less able to engage in social distancing measures, and are less able to work from home and self-isolate when required (2). Emerging evidence from the Mental Health Foundation’s Mental Health in the Pandemic study also indicates that a higher proportion of members of BAME communities are experiencing financial concerns, fear and anxiety than members of the non-BAME population. Furthermore, people from BAME communities are more likely to be in precarious work and where furloughing may not have been offered (12). Understanding and supporting adherence to mitigation measures among this population is therefore critical.

The aim of this study was to gain a better understanding of how people from low-income and BAME communities are adhering to social distancing and self-isolation measures during the COVID-19 pandemic, and to explore in detail the reasons underpinning this behavior.

## Methods

Participants over the age of 18 years from BAME, and low-income White backgrounds were recruited via social media channels. We chose to sample these communities because we considered that their views might differ from and be under-represented in previous studies, and that they might face particular challenges to adherence (e.g. due to financial or family circumstances). At the time of interviews, prevalence of COVID-19 was also greater among these groups, as well as evidence of poorer outcomes among those who contracted the virus (13).

In the first instance, we approached existing contacts with community groups and invited them to share our recruitment advertisement on their social media channels. Using a snowball approach, we asked community group leaders to suggest other group leaders who may be willing to share the advertisement on social media. We invited interested individuals to contact the research team via email. Potential participants were sent an information sheet about the study. All interviews were conducted via the telephone or using the online platform Zoom. Audio recorded verbal consent was obtained.

Interviews were conducted between the 8^th^ and 31^st^ July 2020 and lasted between 21 to 55 minutes. At the time of interviews, non-essential shops and places of worship were allowed to open, and in England, groups of six were allowed to meet outside. People were still required to stay two meters apart, and isolate if they, or their household, experienced symptoms of COVID-19. Face masks were compulsory on public transport, and from the 24^th^ July were also mandatory in shops. Participants were asked about their understanding and perceptions of government mitigation measures, and their decisions about social distancing and self-isolation behavior during lockdown (Supplement 1).

Following the stages of thematic analysis (14), two researchers independently read transcripts and assigned initial codes to the data. Possible themes relating to participants understanding and perceptions of Government guidance for reducing transmission of COVID-19, and their experiences and behaviors during the pandemic were identified and refined through discussion. Data were checked against an initial framework and refinements made as necessary. For each theme in the framework, charts were developed, and relevant text copied verbatim. Charts were used to identify common concepts within and between participants, and explanations sought for divergence. Participants were given the opportunity to discuss the analysis and interpretations with the researcher team via Zoom or telephone meetings. Two participants engaged in these discussions, contemplating how their behavior fit with the identified categories of adherence, and providing feedback on the final themes.

## Results

A total of 20 participants (13 female) took part in the interviews. Participants were between the ages of 18 and 65 years and from Black African and Black Caribbean (N=4), Asian (9) and White (N=7) ethnic groups. The average (mean) Index of Multiple Deprivation decile was 4.15. Four participants reported that they had had COVID-19, or symptoms of COVID-19 in the household.

### Results of the thematic analysis

Thematic analysis showed that participants engaged in active evaluation of infection risk and control measures, following which three context-specific patterns of adherence were identified (Figure 1).

**Figure 1:**
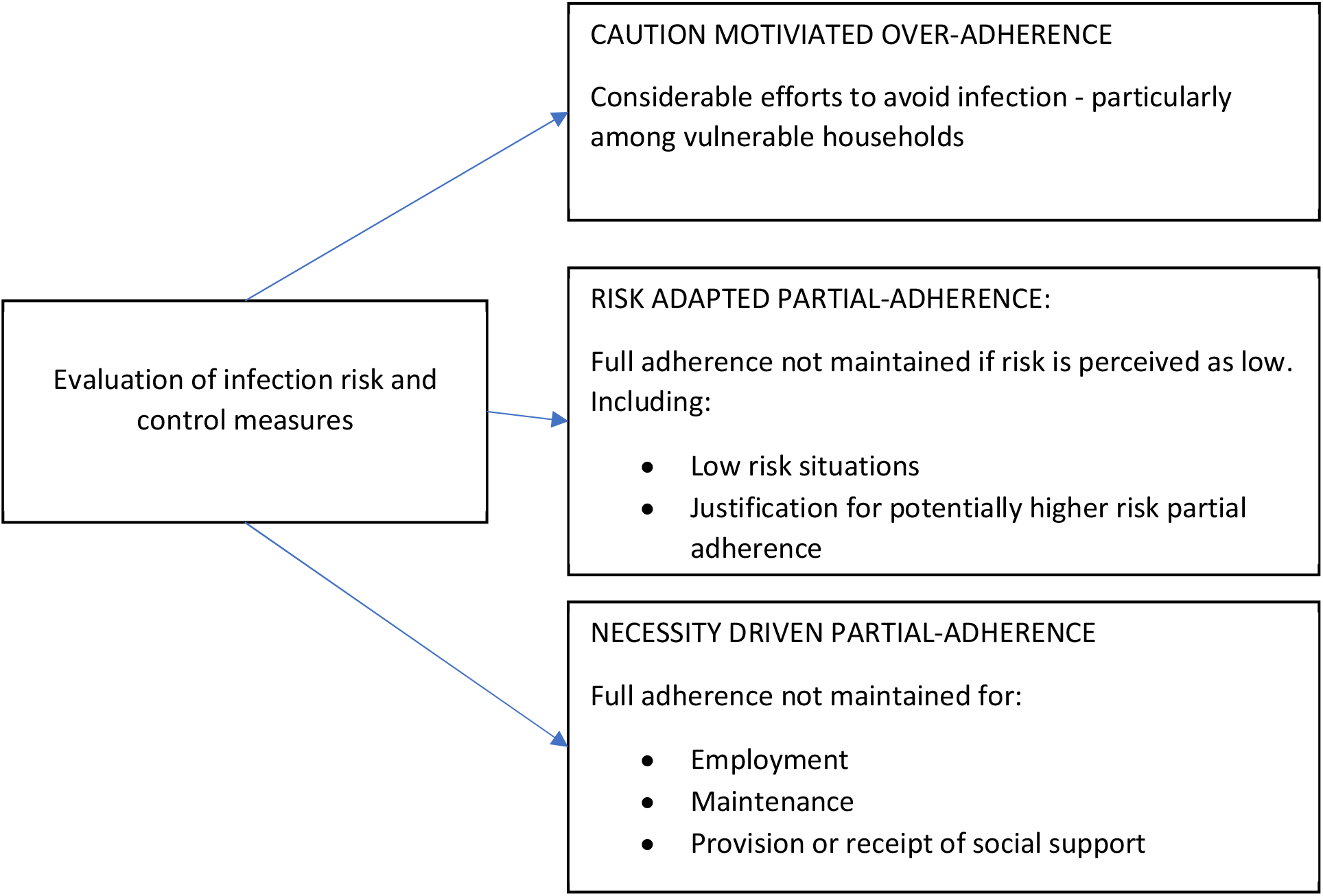
Patterns of adherence to social distanicng and self-isolation measures during the COVID-19 pandemic

### Evaluation of infection risk and control measures

Although participants were eager to understand how they could protect themselves, their households, and their communities, they often struggled to keep up to date with and make sense of the ‘constantly changing’ advice and recommendations (Table 1 – Quote 1). There was evidence of a lack of understanding of critical terms, such as self-isolation, and how to self-isolate safely (Quote 2). Participants were often unclear as to whether they should be isolating (Quote 3), or shielding (Quote 4), and felt uniformed about the number of cases of COVID-19 in their local areas (Quote 5).

**Table 1:** Evaluation of infection risk and control measures

|  |  |
| --- | --- |
| Quote 1 | *"I think the speed at which things were changing was quite confusing. One day there would be one set of principles and then the next day it was all completely changed. It was hard to keep up" (Female, White)* |
| Quote 2 | *"I always found that quite confusing about who should isolate. The guidance around them, and how many days" (Participant 20, White Male).* |
| Quote 3 | *"Um there were a couple of times we may have been displaying symptoms, but we knew, we hadn't isolated for one of those times in fact, to be honest. I guess we took, I would say a vaguely educated risk because no one knew what was happening really" (Participant 03, BAME Male)* |
| Quote 4 | *"I've got high blood pressure and so then that... in some places, in some of the reports it put me at risk. Then some it didn't" (Participant 01, BAME Female).* |
| Quote 5 | *"You want to understand how many people are affected in ((region)). How many deaths. And so on, and so forth. So, you can start to form your opinions" (Participant 08, BAME Male).* |
| Quote 6 | *"I think people could interpret it the way that they wanted to" (Participant 14, White Female).* |
| Quote 7 & 8 | *"You can't trust everything a government says... we have to use our common sense and other things to survive" (Participant 11, BAME Female).*<br>*"It's down to me to decide how I interpret information and take it on board" (Participant 03, BAME Male)* |
| Quote 9 | *"You've got to look after yourself and your family and do with is right for yourself and your family and you won't go far wrong" (Participant 13, White Female).* |

Due to a lack of scientific certainty surrounding the guidance, messages were often considered to be ‘open to interpretation’ (Quote 6). Participants described a need to decide for themselves how to respond to the information they were receiving, and to use their judgement, rather than simply following the guidance (Quote 7 and 8). Decisions appeared to be made on the basis of their individual situations and circumstances, putting the health and needs of their family ahead of government guidance (Quote 9).

### Caution motivated super adherence

Participants described a willingness to adhere to mitigation measures in order to protect themselves and their households from the virus, or to protect others around them. Those who considered themselves to be vulnerable, or at high risk, appeared to be particularly willing to engage in mitigation measures. This was particularly true at the start of the pandemic when anxieties were high (Table 2 – Quote 1). Participants explained how it was important for them to adhere to the restrictions in order to protect their communities, and one participant who had experienced symptoms of COVID-19 described how she was willing to isolate to protect those around her (Quote 2).

**Table 2:** Caution motivated super adherence

|  |  |
| --- | --- |
| Quote 1 | *"I did obey because I don't want to get sick and drop dead" (Participant 01, BAME Female)* |
| Quote 2 | *"But we do have to be selfless and just [adhere to the restrictions] for the rest of your community and the nation" (Participant 02, BAME Female).* |
| Quote 3 | *"Now we're extremely careful here. Being of Asian origin myself I know there's more chances of me catching COVID like Asian and African people than your – compared to Caucasian people, so I'm being extra careful" (Participant 12, BAME Male).* |
| Quote 4 | *"I think when the news came in around the death rate was increasing, I decided after two weeks or a week not to go shopping at all and avoid the supermarket" (Participant 05, BAME Male).* |
| Quote 5 | *"I don't feel protected by the government. I felt the easing of the lockdown felt a bit early. I carried on exactly the same for a while" (Participant 15, White Female).* |

Over time, knowledge of the virus increased, and it became apparent that certain groups were particularly vulnerable. Increased efforts were taken by participants from vulnerable groups to protect themselves (Quote 3) or vulnerable members of their households. Indeed, many of the participants who considered themselves or their households to be vulnerable, and felt that the risk of exposure to the virus was high, reported engaging in additional, precautionary measures to protect themselves and their families (Quote 4). Super-adherence was particularly prominent among vulnerable participants who did not consider government recommendations to be sufficient for protecting them (Quote 5).

### Risk-adapted partial-adherence

A second pattern of adherence included breaking lockdown rules if it was perceived as safe to do so (Table 3). Partially-adherent behaviors were justified by participants, either because they were genuinely perceived as being low risk or because there were sufficient inconsistencies in key messages to allow participants to present their behavior as low risk. Low risk behaviors included those that were considered unlikely to contribute to the transmission of COVID-19, as well as behaviors that were considered safe for the individual participant. Behaviors that were considered unlikely to contribute to the transmission of COVID-19 included those that did not result in close contact with others. For example, one participant described a willingness to leave the house on more than one occasion, a behavior that was not permitted at the time, because he considered it very unlikely that he would come into contact with others (Quote 1). Behaviors that were considered safe for the individual were usually based on individual perceptions of risk. For example, those who did not consider themselves to be at high risk described how this had made them less inclined to adhere to social distancing guidelines (Quote 2 and 3), whilst potentially overlooking the risk to others.

**Table 3:**
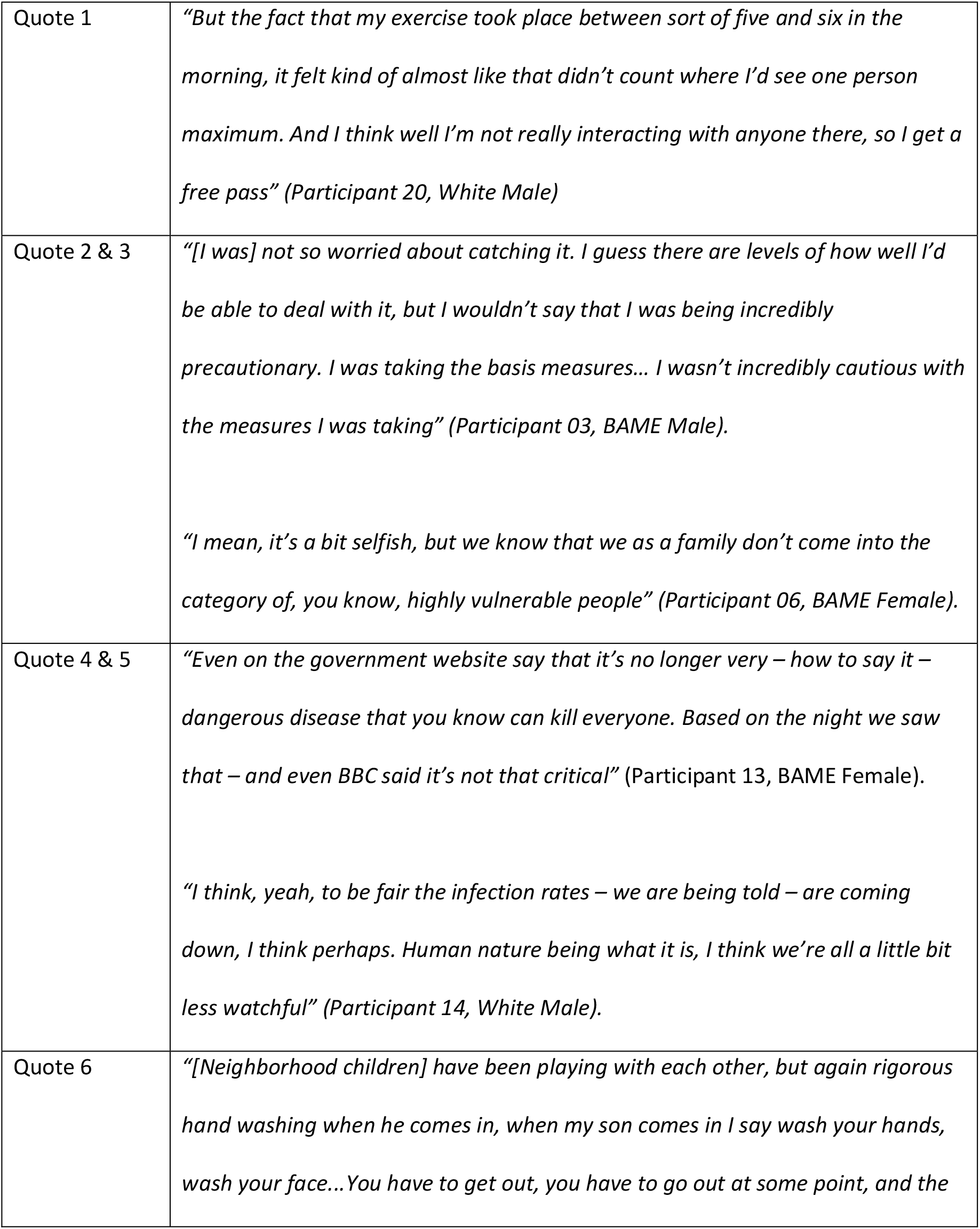
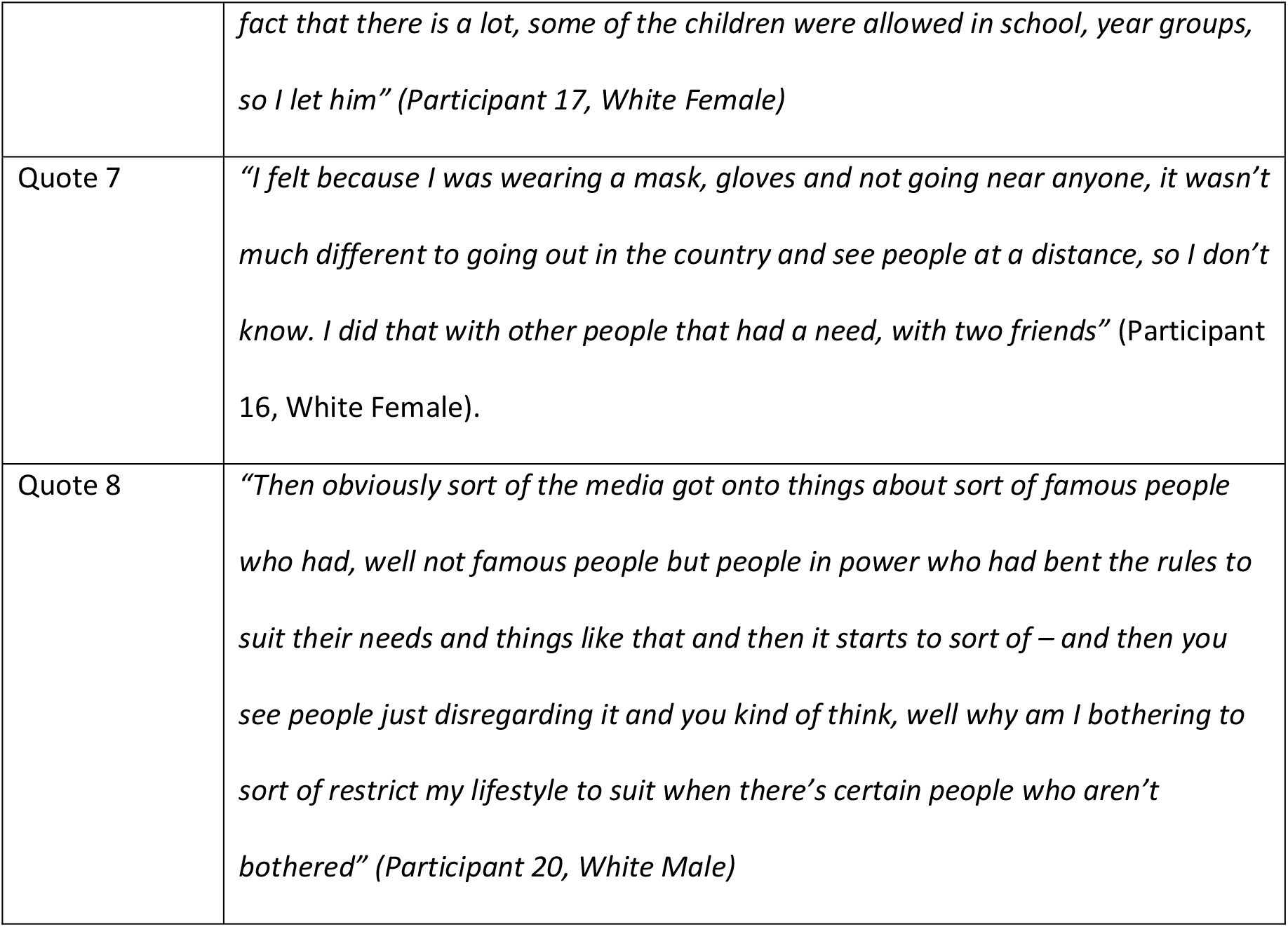
Risk adapted partial adherence

Any apparent ambiguities or inconsistencies in information could be used to justify partial-adherence (Quote 4 & 5). One participants explained how she had allowed her son to play with his friends as key worker children were allowed to remain in school (Quote 6). Another participant was willing to meet a friend during lockdown because she reasoned that it was no different from seeing others’ outside at a distance (Quote 7). A third participant, who described himself as low risk, outlined how breaches of lockdown among influential figures reduced his willingness to restrict his own lifestyle (Quote 8). Although these behaviors may not have been low risk, there was sufficient uncertainty to allow participants to justify breaking the rules. This appeared to be a post-hoc justification for rule breaking; resulting from observations of the behavior of others.

### Necessity-driven partial-adherence

Participants described situations in which they felt they had no choice but to break the rules around social distancing or self-isolation. Participants described a need to find a balance between staying safe and maintaining their mental health and wellbeing (Table 4: Quote 1), or to continue to work because of financial concerns and responsibilities (Quote 2). Indeed, there was perceived pressure from those in management positions to return to work – even when it was against official advice (Quote 3). In another case, a participant who lived in rented accommodation felt under pressure to allow her landlord to enter her home for maintenance purposes, even though she did not feel comfortable allowing him to do so (Quote 4). Other necessities included for religious purposes, as one participant described how members of her household had broken social distancing rules to attend Church services or for Bible studies (Quote 5 and 6). One participant described a need to break social distancing rules following the death of her farther, and later to provide support for bereavement (Quote 7).

**Table 4:** Necessity-driven partial-adherence

|  |  |
| --- | --- |
| Quote 1 | *“Initially there was a lot of worry, it’s still obviously taking precautions, but now it’s kind of trying to find the balance between making sure you’re sticking to the guidelines and being extra safe while still trying to maintain your wellbeing and social interaction, and that side of things” (Participant 10, BAME Female).* |
| Quote 2 | *“They might have just a cold or a sniffle and they would come in [to work]. I know that because they’re not on a big wage, and I know you can get the sickness and all the rest of it, but it’s not going to help when you’ve got a mortgage to pay” (Participant 18, White Female).* |
| Quote 3 | *“I had to narrate to my manager when he was trying to say ‘Why are you not coming in?’... it was into lockdown, April or May, and I was coming out of my illness... I’ve got the emails to prove it... I had to narrate. I had to narrate the government guidelines to stay at home” (Participant 04, BAME Female).* |
| Quote 4 | *“Even if he was not allowed to come in because everyone was not – it was during the first month of the lockdown. No-one was allowed to go [out of the] home except for food and exercise. He came twice to have a look at the house. I don’t know” (Participant 07, BAME Female).* |
| Quote 5 & 6 | *“I mean he [participant’s son] went to church because... well he felt he had to” (Participant 08, BAME Female).*<br>*“She’s got her church friends that goes round. But I think they just sit in the garden or sit in the kitchen, the two of them and do their bible studies” (Participant 08, BAME Female).* |
| Quote 7 | *"I have been to see my step-mum, obviously, after the loss of my father"*<br>(Participant 17, White Female). |
| Quote 8 | *"I did actually realise I was in a bit of a state. I did go and have a distance walk with one of my sons and just basically talked"* (Participant 16, White Female) |
| Quote 9 | *"I did then meet up with another friend who was finding lockdown difficult, which we weren't supposed to at that point. We met at a distance on a walk. I felt mentally I needed that"* (Participant 16, White Female). |
| Quote 10 | *"They've gone out for the last two months. They started going out to do exercises and go and meet up with friends"* (Participant 04, BAME Female). |

A common motive for breaking social distancing guidance was for the sake of their own mental health and wellbeing, or that of their friends and family. One participant described how she had met with her son during lockdown as she was struggling with her anxiety (Quote 8). This participant later described going out during the lockdown period to meet a friend who was also struggling to cope with social distancing measures. Social contact with anyone outside the household was prohibited at the time (Quote 9). One parent was particularly anxious about the mental wellbeing of her children, and described how she had allowed her children to meet with friends before lockdown restrictions were lifted because she felt that they had needed it (Quote 10)

## Discussion

### Main findings

Growing evidence highlights the substantial impact of the lockdown measures on individuals from BAME communities and those on low income (13), with these individuals facing additional barriers to adherence to government imposed mitigation measures (2). In line with previous research, participants in the current study reported engaging in behaviors that were not always in line with the government’s social distancing and self-isolation advice. However, these acts of partial-adherence were not always high risk or avoidable. We suggest that participants made risk-adapted decisions based on their perceptions of the degree of transmission risk entailed by the behavior (for themselves and others), in relation to the importance of the activities they wanted or needed to undertake. We outline three context specific patterns of adherence: 1) caution-motivated super-adherence 2) risk-adapted partial-adherence and 3) necessity-driven partial-adherence. Our findings highlight the need for different forms of intervention, as well as additional research into the impact and risks associated with these patterns of adherence.

The uncertainty surrounding the etiology, transmission and consequences of the virus, and the effectiveness of mitigation measures resulted in a situation in which scientific uncertainty and misinformation was common (15, 16). Government guidance and information changed rapidly in response to emerging evidence and advice. This led to increased confusion, and scepticism among the public regarding the best way to protect themselves, their families and their communities from the virus, and from the negative impact of mitigation strategies. This loss of credibility in government official advice led participants to interpret and act upon the information they were receiving in ways that suited their needs (10). Participants appeared to systematically assess their level of vulnerability, exposure, and ability to mitigate risk, and weigh that against the need for and implications of the behavior. This active decision-making process appeared to inform subsequent behaviour, which did not always conform fully with government advice.

One pattern of adherence involved participants engaging in measures that were additional to those recommended by the government, and continuing to adhere to strict social distancing guidance after restrictions were lifted. Participants who followed this pattern of adherence were concerned about their own vulnerability, or that of household members, as well as about the effectiveness of the mitigation measures in place at the time. A small number of participants in the current sample were able to continue with more stringent social distancing measures in the short term, and it is critical that support is available for vulnerable individuals from BAME communities and those on low income to be able to maintain this level of adherence when necessary. As much research highlights that social distancing is particularly problematic among these individuals (2), support is urgently required for vulnerable individuals who need to engage in additional protective measures when government-imposed restrictions are lifted. However, as considerable research now notes the detrimental impact of lockdown measures on wellbeing, it is also important that people feel confident to safely interact with others once measures are lifted. Indeed, in accordance with previous research (17) nearly all participants described the detrimental impact of mitigation measures on their, or their family’s mental health. This may be particularly problematic for BAME communities, as there is growing evidence that this population are experiencing greater financial worries, fear and anxiety than the non-BAME population (12). Sustained social distancing due to fear and anxiety is likely to have a substantial negative impact on wellbeing over time (10, 18).

A second pattern of adherence involved infringing rules around social distancing if it was perceived as safe to do so due to risk of transmission of COVID-19 being low (risk-adapted partial-adherence). Participants in the current study described leaving the home for physical activity on multiple occasions, or to meet with others at a safe distance outside the home (behaviors that were not permitted at the time). These behaviors were justified by participants as they were viewed as low risk; either because the participant would not be in close contact with others, or because alternative methods of protection (hand hygiene, face coverings) were used. Classifying this behavior as ‘risky’ in the same way as those who engage in high risk (e.g., indoor) contact may be unhelpful, particularly as physical activity is likely to have a positive impact on wellbeing (19, 20). With essential social distancing measures in place this form of partial-adherence could potentially be a lower risk way of obtaining much needed social support. Provision of information to allow individuals to make informed decisions about the risk associated with particular behaviors, places or situations may be beneficial. This information could support people to identify and avoid situations in which the risk of transmission to vulnerable individuals is high, and facilitate appropriate use of infection control measures when in lower risk situations.

In some situations, participants justified breaking social distancing rules because they did not consider themselves or their household to be vulnerable. The potential impact on transmission of COVID-19 beyond their household did not appear to have been considered. Other participants used comparisons to other situations in order to provide justification for their partial-adherence, such as key worker children being allowed to remain in school. However, the extent to which these behaviors or situations are genuinely low risk may be questioned. Indeed some participants, particularly those who considered themselves to be of low risk, appeared to be using information selectively to justify ignoring difficult social distancing rules. The cumulative impact of small acts of non-adherence is still unknown (21). Whilst the individual breaking lockdown may consider themselves to be at low risk of the more serious consequences of COVID-19, the aim of the lockdown was to reduce contact between people to reduce the burden of disease overall in the population. Additional research is needed to understand the true impact of risk adapted partial-adherence on transmission of COVID-19. Highlighting the potential societal impact of small acts of non-adherence in this population may also be beneficial.

A final pattern of adherence involved engaging in potentially risky behavior due to a perceived need or pressure (necessity-driven partial-adherence). This could be motivated by a need to maintain the mental health and wellbeing of themselves or others, to continue to work and earn a living, to deal with emergencies, or for religious reasons. These participants perceived a critical need to engage in these behaviors and felt that they had no choice in the matter. These pressures are likely to be greater among individuals from BAME communities and those on a low-income, who are less able to work remotely or adhere to social distancing and self-isolation guidance (12, 13). In order to improve engagement with lockdown measures, it is crucial that financial, tangible and social support is available. Indeed, previous research highlights that adherence increases when isolating individuals are financially compensated (1) and receive the help and support they need (3). Working with management teams to prevent pressure being placed on vulnerable staff members is also essential.

### Strengths and limitations

Although every effort was made to recruit a diverse and representative sample, we acknowledge that our use of social media may have resulted in a biased sample. Much of our recruitment was via COVID-19 support pages, and previous research has shown that use of social media during the pandemic is associated with increased levels of anxiety (22) and misinformation (23). It is therefore possible that our sample of volunteers are not representative of those who do not use social media for COVID-19 related support or information. Likewise, participants did not necessarily have symptoms of COVID-19, and were therefore discussing breaches of social distancing, rather than self-isolation. Responses may have been different among a population who had experienced symptoms of COVID-19.

Our study may also have been influenced by response bias. It is possible that participants were unable to accurately recall attitudes and behaviors at the start of the pandemic, or did not feel able to disclose risky or substantial breaches of lockdown to the research team. Although participants in the sample were willing to share examples of partially-adherent behavior, they may not have been willing to share experiences of more risky behavior during the interviews.

Finally, interviews were conducted in July 2020. During this period, lockdown measures were being eased, and cases COVID-19 were falling. Alongside changing rules and guidance, knowledge, attitudes and behavior also change rapidly. Attempts to transfer the results of this study to other populations, or periods of lockdown must be made with caution.

Despite limitations, this study revealed a need to consider different patterns of adherence among BAME individuals and those on low-income. Further research is now needed to explore the potential impact of social and cultural diversity between the interview participants in more detail, as well as the transmission risks associated with the different patterns of adherence to social distancing and self-isolation.

### Implications

Our research has identified different patterns of adherence among BAME individuals and those on low income, each with different associated implications and risks. Whilst previous research depicts an overall lack of adherence to mitigation measures, we highlight that there are at least three patterns of adherence, and different forms of intervention will be needed to support individuals to protect themselves, their households and their communities from COVID-19 and the imposed mitigation measures. This may include provision of practical and social support for those who need it. Information for those who over-estimate their own personal risk (super-adherence), and for those who underestimate the societal impact of non-adherence may also be beneficial. Further research is needed to explore the impact and risks associated with categories of adherence, including the cumulative impact of small episodes of non-adherence at a population level.

### Conclusions

Although participants reported partially-adherent behavior, this was the result of a complex decision making process regarding the risks and benefits of engaging in the behavior, often with clear attempts to reduce risk as much as possible. Participants appeared to actively make decisions to engage in behaviors that they considered to be safe and/or necessary, leading to three patterns of adherence. Provision of practical and financial support could reduce the number of people who feel that they have to break lockdown rules, and provision of clear information could ensure that people are able to assess the risk associated with engaging in particular behavior.

## Data Availability

The datasets used and analysed during the current study are available from the corresponding author on reasonable request.

## Declarations

### Ethics approval and consent to participate

Ethical approval was provided by the NHS Health Research Authority London – Queen Square Research Ethics Committee (20/HRA/2549). All interview participants verbally consented to take part in the study.

### Consent for publication

All participants provided verbal or written consent for data to be included in publications.

### Availability of data and materials

The datasets used and/or analysed during the current study are available from the corresponding author on reasonable request.

### Competing interests

None declared

### Funding

This study was funded by the National Institute of Health Research (NIHR) Health Protection Research Unit in Behavioural Science and Evaluation at the University of Bristol, in partnership with Public Health England (PHE) and by UK Research and Innovation (UKRI) / Department of Health and Social Care (DHSC) COVID-19 Rapid Response Call 2 (grant number MC_PC 19071).

The views expressed are those of the authors and not necessarily those of the NIHR, the Department of Health and Social Care, or PHE. The funders had no role in the design of the study, collection, analysis, and interpretation of the data, or in writing the manuscript.

### Authors’ contributions

Conceived the study: All authors

Study design: All authors

Analysed the data: SD, KM, LY

Interpreted the data: All authors

Drafted the manuscript: SD

Reviewed the manuscript and approved content: All authors

Met authorship criteria: All authors

## Acknowledgements

Lucy Yardley is an NIHR Senior Investigator and her research programme is partly supported by NIHR Applied Research Collaboration (ARC)-West, NIHR Health Protection Research Unit (HPRU) in Behavioural Science and Evaluation, and the NIHR Southampton Biomedical Research Centre (BRC).

Sarah Denford, Gemma Lasseter, Mathew Hickman, Isabel Oliver, and Helen Lambert are supported by the NIHR Health Protection Research Unit (HPRU) in Behavioural Science and Evaluation at the University of Bristol in partnership with Public Health England.

Louise Smith and James Rubin are supported by the NIHR HPRU in Emergency Preparedness and Response at King’s College London in partnership with Public Health England.

Charlotte Robin is affiliated to the National Institute for Health Research Health Protection Research Unit (NIHR HPRU) in Emerging and Zoonotic Infections at University of Liverpool in partnership with Public Health England (PHE), in collaboration with Liverpool School of Tropical Medicine and The University of Oxford, the NIHR HPRU in Gastrointestinal Infections at University of Liverpool in partnership with PHE, in collaboration with University of Warwick and the NIHR HPRU in Behavioural Science and Evaluation at University of Bristol, in partnership with PHE. Charlotte Robin is based at PHE.

## Notes

### Competing Interest Statement

The authors have declared no competing interest.

### Author Declarations

Ethical approval was provided by the NHS Health Research Authority London Queen Square Research Ethics Committee (20/HRA/2549). All interview participants verbally consented to take part in the study.

